# Visual field improvement in neglect after virtual reality intervention: a case study

**DOI:** 10.1101/2021.05.04.21256247

**Authors:** Michael Christian Leitner, Stefan Hawelka

## Abstract

**Objective:** Studies on neuropsychological rehabilitation of visual field defects provide an inconsistent picture regarding the effectiveness of so-called “restorative approaches” in visual field recovery (VFR). During a current research project on the clinical evaluation of VFR - in combination with head mounted virtual reality displays (HMD) - a patient (“Patient 7”) suffering from visual neglect was investigated. Although the concept of VFR is originally not intended for patients suffering from *higher* cortical regions (as in neglect), we hypothesized that due to the strong attention-demanding training situation in HMDs, neglect patients might benefit from these intervention procedures based on restorative approaches.

**Methods and Analysis:** Patient 7 was examined perimetrically using a “Humphrey Field Analyzer”, “Goldmann Perimetry” and our newly developed and validated eye-tracking supported perimetric methodology “Eye tracking based visual field analysis” (EFA). Based on these high resolution results from the EFA, the exact location of the transition area between intact and defect visual field of Patient 7 was assessed. Next, bright light stimuli were placed along this area in our newly developed HMD “Salzburg Visual Field Trainer” (SVFT). The aim was to stimulate neuroplasticity - according to the concept of restitutive approaches - in the corresponding cortical areas of the patient. Patient 7 trained with the SVFT for a time period of 254 days. In 6 appointments the objective and subjective rehabilitation progress was assessed.

**Results:** Perimetric assessment with the EFA shows an expansion of Patient 7’s visual field of 48.8% (left eye) and 36.8% (right eye) after 254 days of training with the SVFT. Individual areas in the patient’s visual field show a visual improvement of approximately 5.5° to 10.5° of visual angle. Subjective self-report of Patient 7 additionally shows improvements in self-evaluation of up to 317% in visual field functionality compared to self-evaluation on the first assessment date.

**Conclusions:** The results from Patient 7 indicate that patients suffering from visual neglect potentially benefit from a neuropsychological intervention with HMD based on the restorative concept of visual field recovery. However, further studies with large case numbers and a focus both on daily-life improvements and on a clear distinction between patients with lesions in earlier and higher cortical areas are needed to make empirically valid and generalizing statements about our findings.

## Introduction

### Scientific background

Worldwide over 13 million new cases of stroke are reported annually, with stroke being the second major reason for disability (Lindsay et al. 2019). Approximately 50% of people suffering from stroke report visual problems, ranging from central visual issues to eye movement and perceptual disorders (Rowe et al. 2019). Besides visual field defects stemming from lesions in early/primary cortical areas, a widespread clinical disorder after stroke or trauma is visuospatial neglect stemming from lesions in higher cortical areas (Berti, Garbarini & Neppi-Modona 2015). (Visual) neglect describes a multicomponent complex disease with neuropsychological issues in the patients’ processing of spatial information, resulting in decreased awareness for the spatial side opposite to the lesioned cerebral area (e.g. Bisiach 1996; Kerkhoff, Rode & Clarke 2021). Literature distinguishes between egocentric and allocentric neglect, with egocentric neglect being the more common form and left-sided neglect occuring twice as frequent than right-sided neglect (Bowen, McKenna & Tallis 1999; Demeyere & Gillebert 2019). Patients suffering from egocentric neglect “[…*] fail to attend to the contralesional side of space (with reference to their own body midline) [while] allocentric neglect is where [patients] fail to attend to the contralesional side of an object in focus*.” (Demeyere & Gillebert 2019, p.491). Studies show prevalence of neglect in older stroke patients (> 60 years) ranging from 15% to 33.5% and 50% of stroke patients of all ages suffer left-sided neglect (Chen et al. 2015; Linden et al. 2005; Ten Brink et al. 2016). Visual field assessment is usually conducted with kinetic perimetry (e.g., “Goldmann Perimetry” [GP]) and/or static perimetry (e.g., “Humphrey Field Analyzer” [HFA]) (Kerkhoff, Rode & Clarke 2021; Kolling et al. 2012).

### Neuropsychological rehabilitation

There are two main approaches for patients suffering from visual defects. “Visual Field Training” (VFT) describes a methodology that helps patients to learn strategies to *compensate* (thus also known as “Compensatory therapies”) for visual field defects by improvement of saccadic search patterns and increasing attention to blind areas. Conversely, “Visual Field Recovery” (VFR) aims to *reactivate / restitute* lesioned areas (in the visual cortex) by repeated light stimulation which - supposedly - leads to neuronal reconnection (thus also known as “Restitution therapies”). While visual field defects originating from stroke or trauma are classified in the scientific literature as “perception deficit”, visuospatial neglect is widely described as “attention deficit” (e.g., Halligan et al. 2003; Lunven & Bartolomeo 2017). Despite these differences in neurological location and neuropsychological functionality, similar rehabilitation routines are conducted in visual field defects and visuospatial neglect with both based in particular on VFT approaches (e.g., Kerkhoff, Rode & Clarke 2021; Umarova et al. 2011). While VFT currently represents the “gold standard” in clinical rehabilitation, the true effect sizes of VFR are subject to discussion in the scientific community for years (e.g., Bouwmeester, Heutink & Lucas 2007; Frolov, Feuerstein & Subramanian 2017). However, researchers agree that the need for visual rehabilitation and new approaches will further increase in the near future. Trauzettel-Klosinski (2011) states that *“suitable rehabilitative measures chosen after the thorough diagnostic evaluation of a visual impairment and analysis of its effects can usually restore reading ability, improve orientation, and thereby enhance the patient’s independence and quality of life. As the demand for visual rehabilitation is increasing, steps will need to be taken to make it more widely available*.*”* (p.871)

### Controversy

VFR is based on the concept of “visual border stimulation”, the activation of “residual vision capacities”, with various proprietary developments from different research groups and companies. What unites all the concepts, however, is the hypothesis that - by stimulating the “border area” between the intact and the damaged visual area - neurons in the topographically corresponding area of the visual cortex can be re-activated (e.g., Sabel et al. 2013). Sabel, Kasten and Kreutz (1997) reported that patients sometimes detect visual stimuli in the border to the intact visual field. The authors further suggest that 15% of remaining, functionally intact neurons are sufficient to stimulate activity in the affected area. This could, in turn, lead to plastic cerebral changes and result in a partial restitution of the visual field (Rosa et al. 2013).

Some studies reported considerable training effects with border stimulation programs indicating neuroplasticity in the visual cortex (Marshall, Chmayssani, O’Brien, Handy & Greenstein 2010; Mueller, Mast & Sabel 2007; Sabel, Kenkel & Kasten 2005; Poggel, Kasten & Sabel 2004; Sabel, Kenkel & Kasten 2004; Mueller et al. 2003; Zihl & von Cranon 1985). Mueller, Mast and Sabel (2007), for example, reported that their training restored up to 17.2% of the formerly blind visual field of their patients. Marshall et al. (2010) reported an average improvement rate of 12.5%.

Other studies, by contrast, did not find (significant) effects (Glisson 2006; Horton 2005; Reinhard et al. 2005; Balliet, Blood & Bach-y-Rita 1985) and hence the effect of vision restoration is still controversially discussed (e.g., Pollock et al. 2019; Sabel & Trauzettel-Klosinski 2005). Frolov et al. (2017), for example, attest that there are *“remaining nagging questions as to the validity of* [published data] *and the clinical benefit*” (p. 40) and Kerkhoff et al. (2021) states that *“the overall evidence is unclear and thus restorative visual field training cannot generally be recommended [*…*]”* (p. 196). Horton (2005) assumed in his review of the available evidence, that those studies which found beneficial effects of VFR training reported overly optimistic outcomes. The reason for this scepticism is the imprecision of the diagnostic instruments which are traditionally used to assess the extent of the visual field loss (e.g., GP and/or HFA). Thus, traditional diagnostics systems have major limitations in reliably assessing the potential effects of VFR: i.) they lack a stringent fixation control ii.) they allow the patient to (unconsciously) resort to compensation strategies (Schreiber et al. 2006) and iii.) - in case of manual perimetry - are prone to a certain degree of subjectivity of the examiner.

Against this background, we developed - in the course of our current project on the effectiveness of therapeutic interventions on visual field defects - an eye tracking based perimetric methodology for highly accurate pre-, post-, and follow-up assessments, i.e., “Eye Tracking Based Visual Field Analysis” (EFA) (Leitner et al. 2021). The EFA is based on static automated perimetry while additionally taking individual eye movements (with the help of an eye tracker) in real time into account and compensating for them. The EFA provides a standard error of measurement (SEM) of 0.44° of visual angle. Based on participants’ individual results, the EFA provides disattenuated correlation (validity) with established perimetric instruments of 1.00. Results from patients suffering from cortical lesions imply that the EFA is applicable for clinical use and pre-post-assessment enabling the finding of small changes (> 0.5°) in patients’ visual fields.

### Virtual reality

Recently, various research groups began developing and studying perimetric and therapeutic procedures for patients suffering from visual field defects based on virtual reality and mobile concepts. The shared vision is to use so called “Head Mounted (Virtual Reality) Devices” (HMD) and/or mobile applications (based on tablets or cell phones) as valid and reliable tools for diagnostic and therapeutic purposes (e.g., Goh et al. 2018; Leitner, Guetlin & Hawelka 2021; Matsumoto et al. 2016; Wroblewski et al. 2014). The goal is to ease and improve both diagnostics and intervention for healthcare professionals and patients alike. Different studies imply that HMD and mobile visual field assessment tools (e.g., implemented on iPads) are indeed an alternative for established, stationary tools. Anderson et al. (2017) showed that *“Detecting rapid visual field progression may be improved using a home-monitoring strategy, even when compliance is imperfect*.*”* (*p*. 1735). For example, Schulz et al. (2017) state that iPad-based threshold perimetry shows “[…*] reasonable sensitivity/specificity [*…]” (p.346) and Nesaratnam et al. (2017) envisage that “[…*] such a test having a role in assessing bed-bound patients in hospital where access to formal visual field testing is difficult, or indeed in rapid testing of visual fields at the bedside [*…]” (*p*. 1). This approach is particularly helpful for stroke patients who are bedridden or immobile (Spofforth, Codina & Bjerre 2017). Sand et al. (2012) additionally emphasize that the focus in daily neurological practice and rehabilitation is mainly on motor symptoms and that visual impairments must receive more consideration in order to improve patients’ independence and quality of life. Because modern HMD or tablets offer various protocols for connectivity (e.g., 5G, 4G, 3G, WIFI, Bluetooth, NFC, etc.), this hardware can be implemented effortlessly in telemedical routines. In this context, Johnson et al. (2017) showed that mobile perimetric applications produce valid and reliable results in areas with limited medical care. This progress represents a major opportunity for people in developing countries or medically poorly served areas to receive orthoptic and neuropsychological care and rehabilitation, prevent degenerative illnesses and monitor therapeutic progress. In general, HMD hardware constitutes an evolution that will continue to develop in stroke and orthoptic research but also in various other academic fields such as in Traumatic Brain Injury, Parkinson disease or Autism over the next few years (e.g., Canning et al. 2020; Cortés-Pérez, Nieto-Escamez & Obrero-Gaitán 2020; Maggio et al. 2019; Qazi & Raza 2020; Spreij et al. 2020).

In this light, we developed a portable virtual reality-based HMD solution for neuropsychological rehabilitation to both *investigate* and *enable* the effectiveness of therapeutic interventions on visual field defects (Leitner, Guetlin & Hawelka, 2021). Results on the reliability and validity of the device indicate that the “Salzburg Visual Field Trainer” (SVFT) is highly accurate in displaying visual stimuli in predefined areas of the users’ visual field, which is a fundamental requirement for the potential effects of rehabilitation approaches such as VFR. Specifically, SVFT provides an overall accuracy of .990 (sensitivity of .982 and specificity of .994). Concurrently, feedback from patients further indicates that neurorehabilitative training with HMD is practical, easy, and comfortable to perform (in contrast to setups based on classical “Head-Chin Rests” solutions).

### Aim

In our current research project, we found indications that patients suffering from neglect might significantly benefit from VFR when training with SVFT. Due to the scope of our research project, we originally excluded patients suffering from visual neglect from our experimental design and concentrated solely on patients suffering from clearly definable defects in *early* visual cortex (e.g., infarct of the arteria cerebri posterior). However, we later decided to include one patient (i.e., “Patient 7”) - who suffers from visual neglect after trauma (see below for details) - as a designated special interest case. We hypothesized that the highly focused and frequently repeated training situation in HMDs - due to its closed design covering both eyes and, in this way, leading to intense confrontation with the training task - could potentially improve attentional mechanisms in neglect patients (Azouvi, Jacquin-Courtois & Luauté 2017; Kerkhoff, Rode & Clarke 2021; Kerkhoff & Schenk 2012). In the following case report we present our findings.

## Methods

### Patient involvement, privacy and ethical considerations

Patient 7 was not directly involved in the design of this study. Her ID number is not known to anyone outside the research group. Written informed consent (to report all amnestic, clinical and diagnostic details in this manuscript) was obtained from Patient 7. The neuropsychological intervention, the ophthalmologic assessment and the concept of the study was approved by the ethics committee of the University of Salzburg. The present study is guided by the fundamental principles of respect for the individual, the right to self determination, and the right to make informed decisions, i.e., informed consent, as noted in the Declaration of Helsinki. The study is registered in the ICMJE-approved registry “German Clinical Trials Register” (DRKS00025205).

### Patient characteristics

Patient 7 is a 40 to 45 years old woman who suffered traumatic brain injury from an accident in 2018 with extensive posterior medial infarct and posterior infarct on the right with midline shift (ICD: I63.4) and posterior medial infarct on the left and right cerebellar infarct in the AICA flow area resulting in a visual field defect (quadrantanopia lower right and lower left) with egocentric visual neglect symptomatic to the lower left side (see Fig. 1).

**Figure 1.**
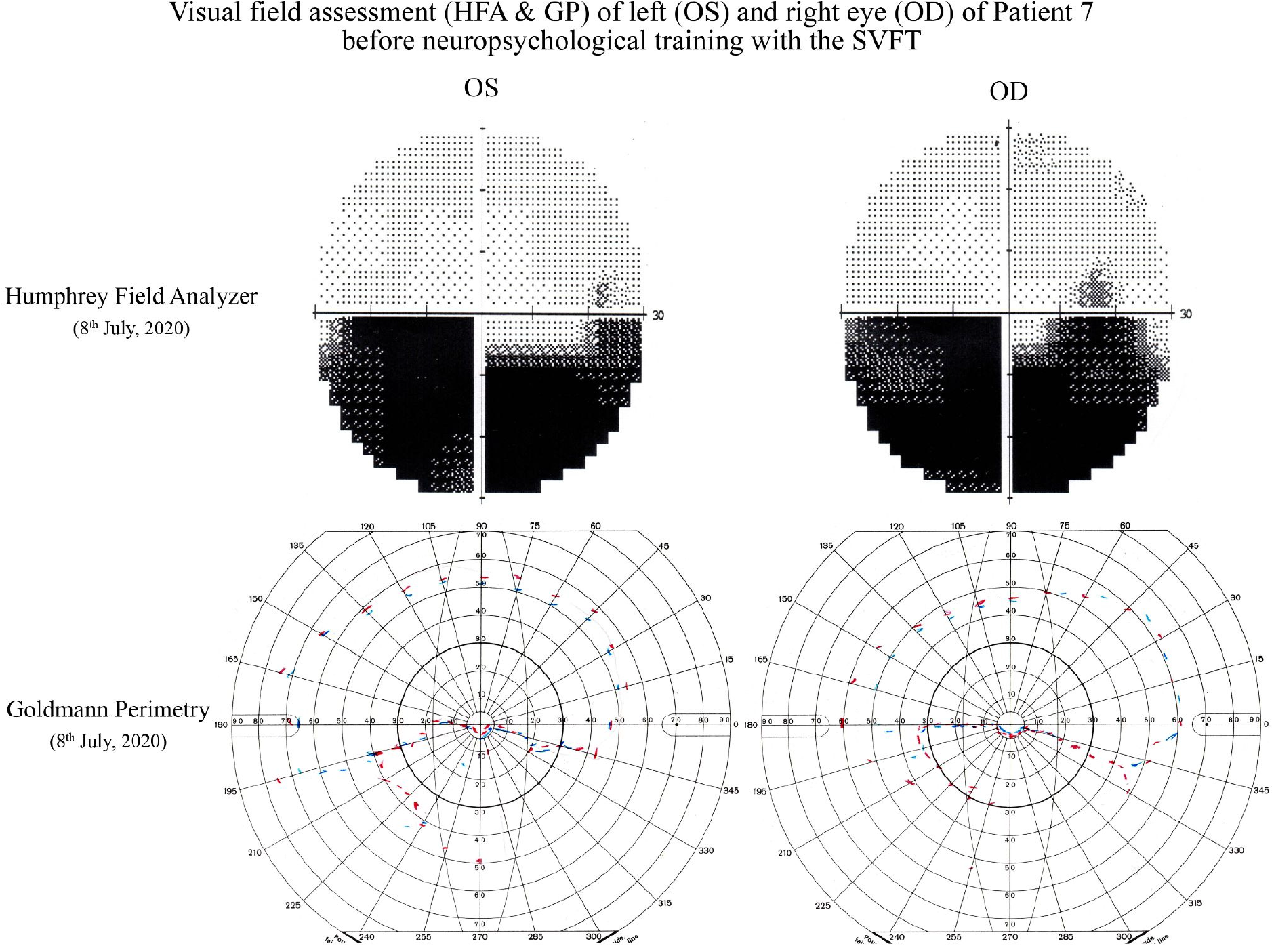
Illustration of visual field assessment of Patient 7 with Humphrey Field Analyzer (HFA) and Goldmann Perimetry (GP) on July 8^th^, 2020. Results indicate a broad visual field defect below the horizontal line. Test statistics from HFA show 10% false positive errors, 11% false negative errors, 1 fixation loss and an assessment duration of 6 minutes for the right eye. For the left eye there are 0% false positive errors, 0% false negative errors, 0 fixation losses and an assessment duration of 5 minutes 28 seconds. Both eyes were assessed with the SITA-Fast algorithm. During visual field testing with GP, Patient 7 was calm, concentrated and acted consistently. Note that blue lines represent Goldmann marker size I and red lines size III, both with a relative intensity of 1.00.

From a neuropsychological perspective Patient 7 is conscious, oriented in time, place and situation. She is unobtrusive in her general perception and attention, her concentration capacity is undisturbed, there are no memory or retentive disorders, and she shows no signs of anosognosia. Her speech production is fluent and well-articulated, her speech comprehension is undisturbed and requests from healthcare professionals are executed promptly and adequately. There are no signs of meningismus, her pupillomotor functionality is undisturbed and pupils are bilaterally isocor and normal light reactive. Patient 7 shows a slightly saccadic gaze sequence, but no gaze paresis or indications of double vision with age adequate accommodation. Patient 7 showed significant difficulties in the “Clock Drawing Test”. Drawing from 12 to 6 o’clock was uneventful, however, the patient did not know what to do next and stopped the test. After some further questioning, the patient first drew 8, 4 and 3 in the left half of the clock face. After repeated questions and assistance, the patient was able to complete the clock. In a second task, the “Line Bisection Test”, Patient 7 showed no significant abnormalities.

### Design, assessment and rehabilitation

The inner 10° of visual field of Patient 7 was diagnosed monocularly in detail with the help of the EFA on July 14^th^, 2020 at the “Centre for Cognitive Neuroscience” of the University of Salzburg. Patient 7’s vision was corrected for acuity during all visual field tests. Based on these diagnostic results the SVFT was configured to present bright light stimuli onto her visual border area, which is between the intact and defect visual field of Patient 7 (see Fig. 2).

**Figure 2.**
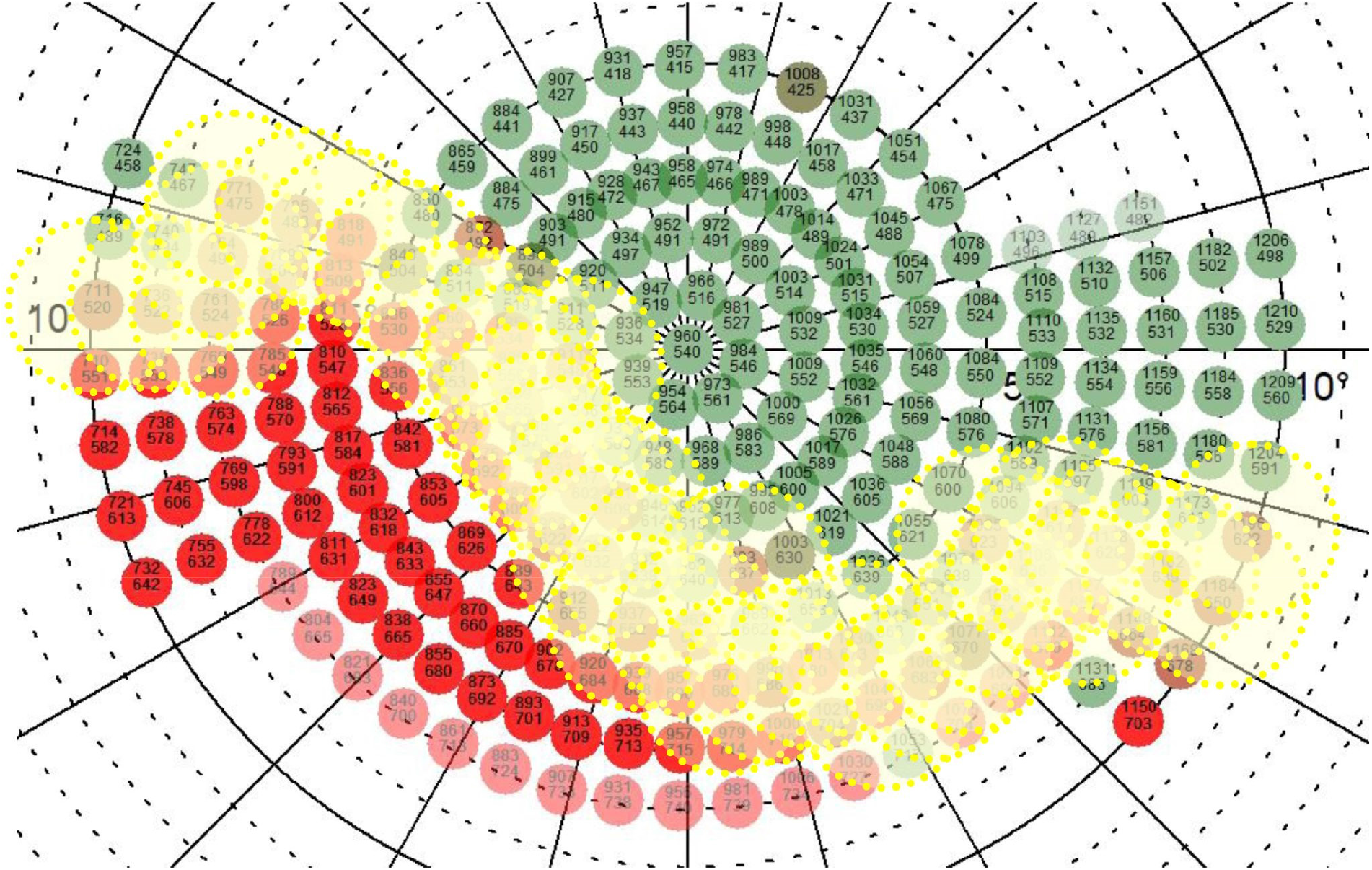
Illustration of placement of 38 training stimuli (yellow dotted circles) along the border area of the intact (green dots) and defect (red dots) inner 10° of the binocular visual field of Patient 7, as assessed with the help of the EFA. The illustration shows training stimuli arrangement after the first visual field assessment with the EFA on July 14^th^, 2020. In a total training duration of 254 days, the visual field of Patient 7 was assessed in regular intervals (M = 51 days (SD = 17)) with the help of the EFA (6 assessments in total). When significant changes in the visual field (> 3°) of Patient 7 after each assessment were found, the training stimuli were rearranged accordingly.

Training stimuli had a size of 3° and a luminance of 1,000 cd/m^2^, appearing for 750 ms in randomized order on a dark background. The presentation of each stimulus was delayed for 2000 ms (plus a randomized duration of up to 1500ms) to give Patient 7 time to react to the displayed stimulus via remote control. This is necessary as the SVFT runs a “stimulus-location adaptation process” during training sessions which is based on user feedback by clicking (or not) the remote control during the delay after every displayed stimulus. If the user detects the stimulus peripherally, its location is moved for 0.8° towards a custom reference point within the central area of the defect visual area. This allows the system to automatically adapt to an improving visual field or small inaccuracies from minor gaze deviations from the fixation cross. On the contrary, if the stimulus is not detected, it is moved away from the reference point for 0.4° towards the center of the patient’s intact visual field, balancing out the therapeutic process in case of inaccuracies, misclicks or loss of already gained training progress. Thus, this behavioral procedure produces a steady approximation of the “real” border across the intact and defect visual field and successively compensates for potential minor gaze inaccuracies of the patient during the training session procedure (Leitner, Guetlin & Hawelka 2021). After 30 minutes of training, the SVFT ends automatically.

Patient 7 was instructed to train twice a day for 6 days a week and extensively informed about the procedure and methodology of VFR and regarding the functionality of the SVFT on the first assessment date (for details see Leitner, Guetlin & Hawelka 2021). At the end of the first appointment, the SVFT was handed over to Patient 7 and an appointment was made for the first interim measurement. The study called for Patient 7 to come into our laboratory at the University of Salzburg approximately every 2 months for visual field testing with the EFA. However, this plan could not be fully adhered to due to the COVID-19 pandemic, which is why some appointments had to be shifted back and forth by a few weeks over the next months. Nonetheless, this led to a mean duration between assessment appointments of 51 days (SD = 17).

### Measures and analysis

Changes in the visual field of Patient 7 are calculated and displayed in two different ways. These are as follows: 1) Percentage change of visual field functionality is based on the ratio of perceived (green dots) and unperceived stimuli (red dots) between the first (0 days) and the last assessment day (254 days), as well between assessment days (0, 64, 105, 182, 217, 254). Areas that were not assessed at the first appointment were not considered for the calculation of indicated percent change - even if these areas were additionally tested in later assessment days due to significant changes in visual field functionalities. As a result, potential improvements tend to be underestimated rather than overestimated. 2) Apparently improved regions in the visual field of Patient 7 are marked in the visual field plots with rectangles. Horizontal and vertical changes of these areas are calculated and reported in degree of visual angle (°). A prerequisite for this calculation is a coherent image of the enhancement of larger contiguous - former blind areas - of at least 3°. Areas that do not qualify for calculation but tend to show apparent improvements are reported but no specific calculation for degree of visual angle is carried out.

In addition to the objective perimetric measurement of potential improvements in the visual field functionality of Patient 7 - which was done with the help of the EFA - we were also especially interested in the subjective valuation of Patient 7 regarding her visual capacities. Thus, before every EFA assessment in our lab, Patient 7 was asked to rate the status of her visual field on a continuum subjectively, ranging from “very bad” to “very good”.

## Results

Visual field test results from the EFA after training for approximately 8 months with the SVFT show an expansion of Patient 7’s visual field of 48.8% (OS) and 36.8% (OD). This translates to a horizontal expansion of approximately 7.25° and vertical expansion of approximately 5.5° of visual angle for the upper left area in OS and to a horizontal expansion of approximately 10.5° and vertical expansion of approximately 5.5° of visual angle for the lower right area in OD (both marked as yellow rectangles in Fig. 3). Improvements marked by the lower right rectangle in OS are too widespread and diffuse to be translated into degree of visual angle, however reporting of overall improvement in this area appears worth noticing.

**Figure 3.**
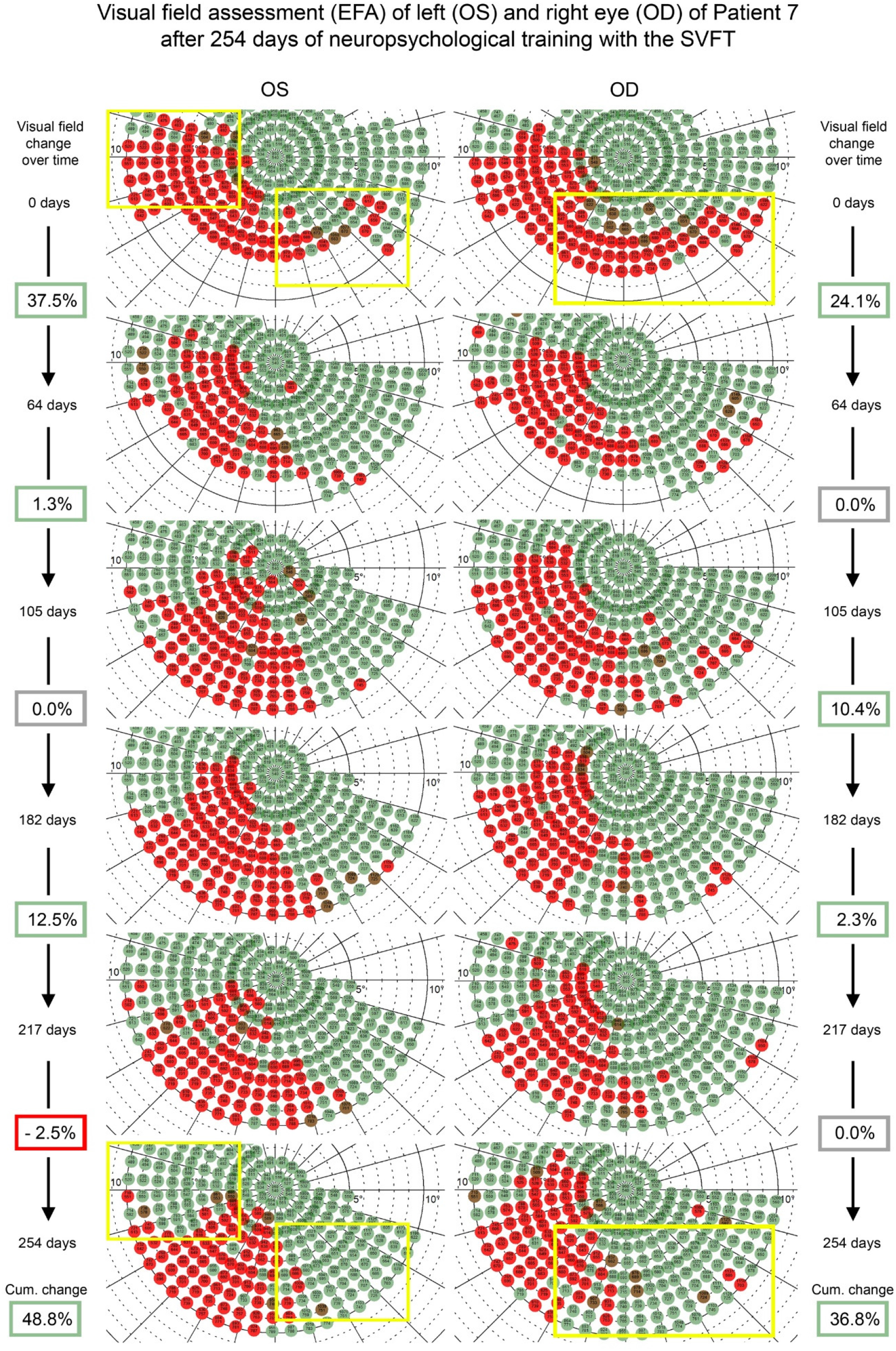
Illustration of visual field tests (inner 10°) - conducted with our perimetric “Eye Tracking Based Visual Field Analysis” (EFA) - after 217 days of therapy with our HMD “Salzburg Visual Field Trainer” (SVFT) of Patient 7 suffering from quadrantanopia and visual neglect symptomatic. Red dots represent stimuli not detected, green dots represent stimuli detected during perimetric assessment. One stimulus stands for approximately 0.75° of visual angle. Note the obvious improvements in stimulus detection - marked by the yellow rectangles - at both sides of the respective visual field defects. Data analysis (by total numbers of undetected stimuli based on baseline assessment) shows total visual field enlargement of 48.8% (OS) and 36.8% (OD) and gradual improvement especially during the first 64 days of therapy (also see Fig. 4). Especially interesting are also the unsystematic improvements in the center of the defect area, which are (naturally) not yet locatable during baseline assessment. Because the EFA repeats undetected stimuli - after displaying a random stimulus that is not recorded - the probability of improvements by random patient’s reaction is highly unlikely. Also, the EFA documents if patients are “trigger happy”, which is not the case with this patient (also see Fig. 4). Dots in dark red represent stimuli not detected the first time but detected the second time. These stimuli usually occur near “visual border areas”, situated between defect and functional visual field areas - as it is observable also in these exemplary results of this Patient 7.

The behavior of Patient 7 during visual field assessment - represented in the EFA’s test statistics - shows no abnormalities, with false negative and false positive rates within usual and similar ranges comparable to standard perimetric analysis of Patient 7 with the HFA (see Fig. 4).

**Figure 4.**
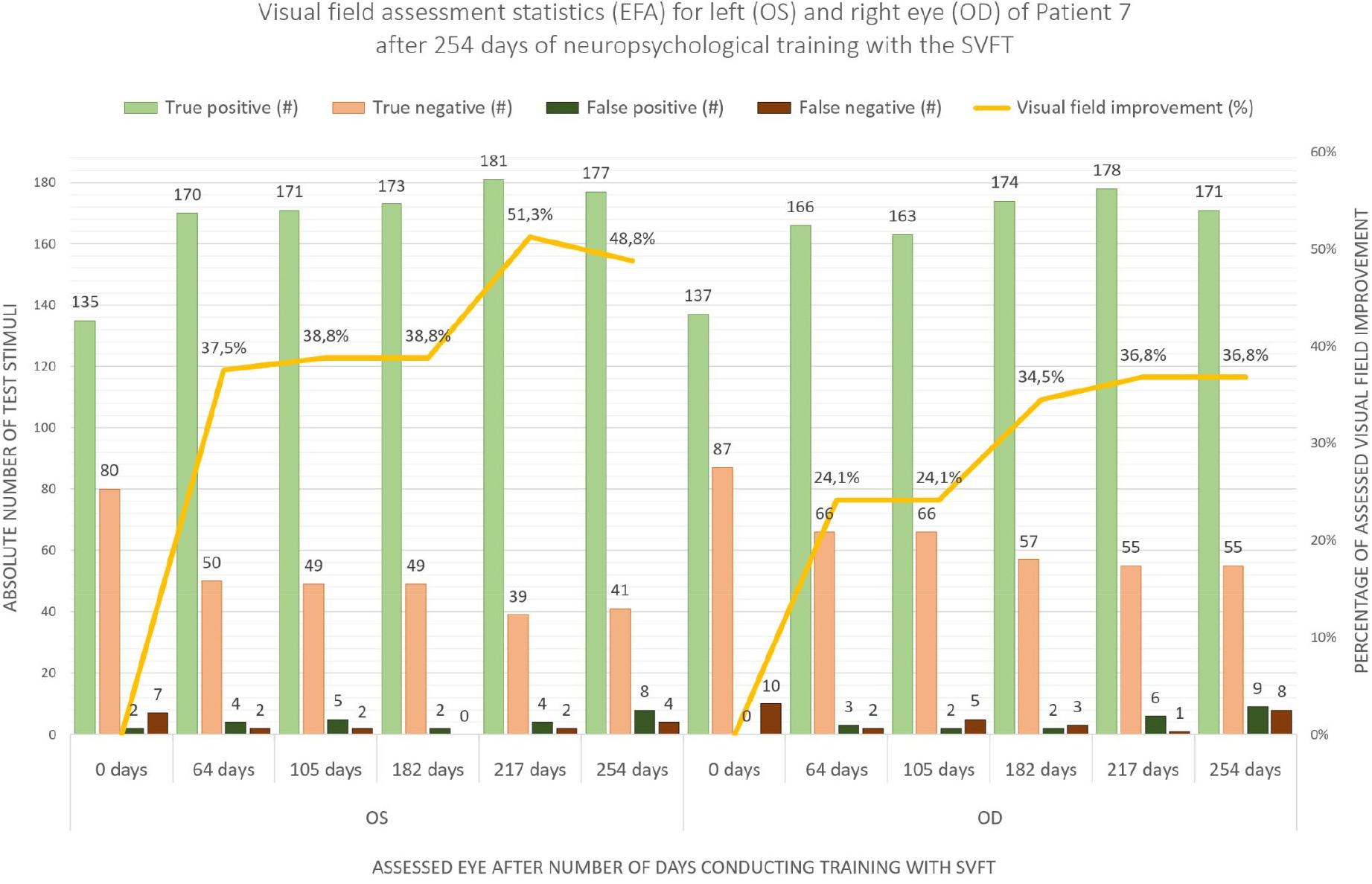
Illustration of EFA’s test statistics of the left eye (OS) and right eye (OD) of Patient 7 after 0, 64, 105, 182, 217 and 254 days of neuropsychological training with the SVFT (x-axis) in absolute numbers of presented test stimuli (left y-axis). Results show no abnormalities, with false negative and false positive rates within similar ranges compared to standard perimetric analysis. The ratio between true positive and true negative stimuli shows improvements in visual functions (yellow line) due to repeated neuropsychological training with the SVFT (right y-axis).

Analysis of the SVFT training log shows that Patient 7 trained consistently at least twice for 30 minutes per day over the entire period. Specifically, Patient 7 performed 136 training sessions within the first 64 days, 88 training sessions within the following 41 days, 171 training sessions within the following 77 days, 104 training sessions within the following 35 days and 111 training sessions within the last 37 days. This results in an average of 2.1 training sessions per day for the first period, 2.1 training sessions per day for the second period, 2.2 training sessions per day for the third period and 3.0 training sessions per day for the fourth and fifth period. This translates to a total mean of 2.5 training sessions per day with the SVFT for the entire period of 254 days.

Additional results show that the perceptual improvements of Patient 7 - because of the training with SVFT - are not only objectively measurable via the EFA, but also reported by the patient herself (see Fig. 5).

**Figure 5.**
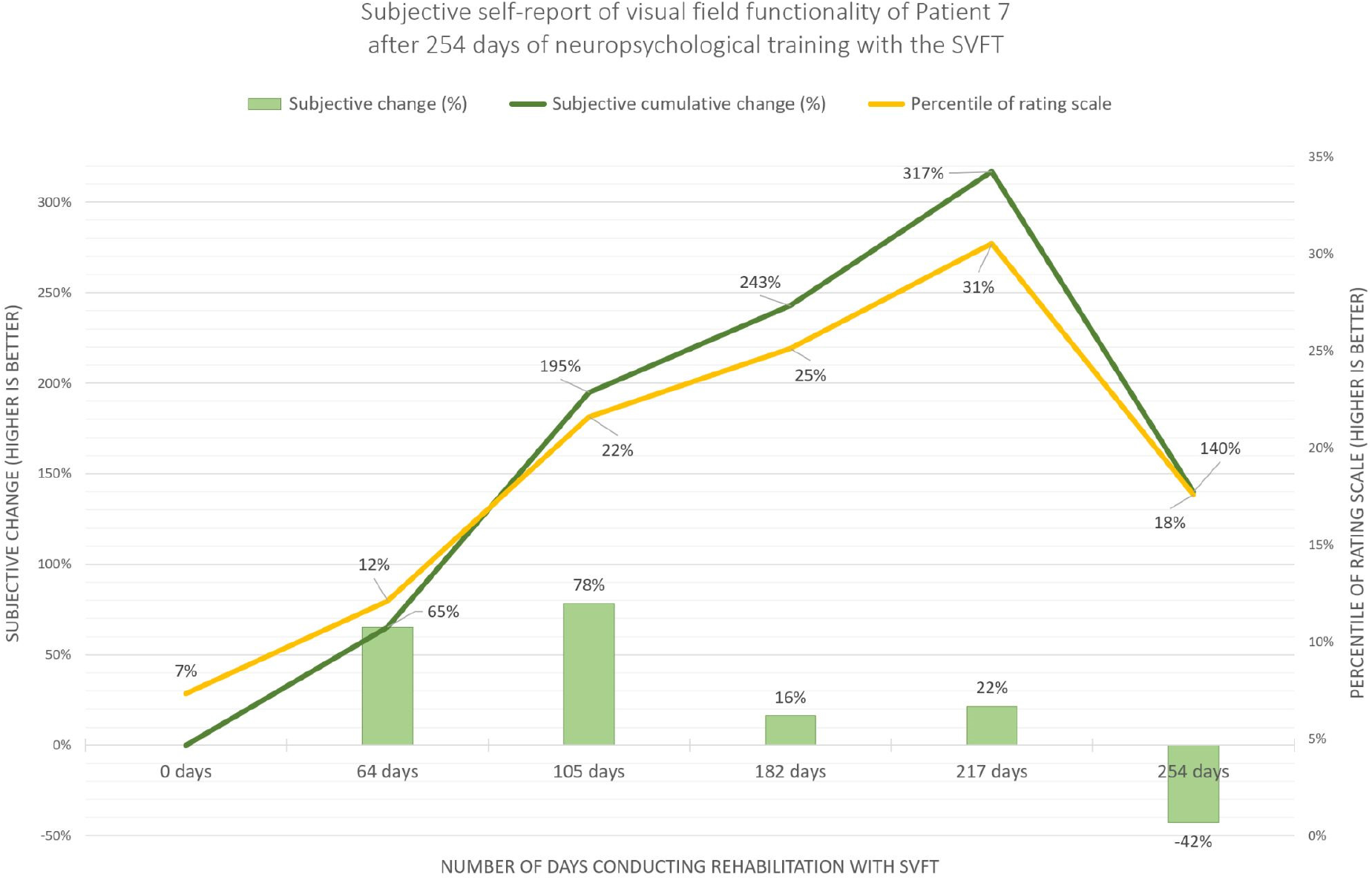
Illustration of subjective self-report (left y-axis) of visual field improvement of Patient 7 after 0, 64, 105, 182, 217 and 254 days of training (x-axis) with the “Salzburg Visual Field Trainer” (SVFT). Results show improvements in self-evaluation (on a continuum (right y-axis) ranging from “very bad” (0%) to “very good” (100%)) of visual field functionality and confirms objective data from repeated perimetric assessment with the EFA. The self-report indicates that Patient 7 feels that she is benefiting from the rehabilitation program and that her visual (-attentional) capabilities improve. We interpret the subjective worsening at the last test time point as a reaction to the preceding announcement of the research team to temporarily stop the therapeutic intervention in order to evaluate the data of the study and to plan further steps. According to her own statements after assessment, the patient would have liked to continue training with the device.

## Discussion

Reliable and conclusive evidence on the effect of neuropsychological interventions to improve visual field defects is scarce and still subject to debate in the scientific community. In our current research project, we aim to test different clinical interventions for their effectiveness and strive to provide new, evidence-based approaches to established methodologies. To create the basic conditions for this we have developed two new tools in the field of diagnosis as well as in the field of rehabilitation. On the one hand, the perimetric tool “Eye tracking based visual field analysis” (EFA) helps us to obtain reliable and accurate measurement results about potential changes in the visual field of patients (Leitner et al. 2021). On the other hand, the neuropsychological tool “Salzburg Visual Field Trainer” (SVFT) creates the virtual reality-based foundation for an accurate, controlled and immersive rehabilitation situation (Leitner, Guetlin & Hawelka 2021). We are currently concentrating on questions regarding “restitutive procedures”, as utilized in so-called “Visual Field Recovery” (VFR) approaches. Since the findings in this context can be described as controversial, we decided to pay strict attention to the neurological background of the respective individual visual field defects when selecting patients. Therefore, we focus mainly on clearly definable “classical” visual field defects that originate in early cortical areas (e.g., stemming from insult of the arteria cerebri posterior) and can be classified as pure “perceptual disorder”. It is our suspicion that earlier studies - besides inaccurate pre-, post-, and follow-up diagnostics, which are due to compensatory eye movements in the course of perimetric examinations - did not elaborate on the exact neurological background of the visual field defects. Specifically, we suspect that i.a., patients with early and higher cortical lesions were mixed in these studies. In addition, it is often difficult and time-consuming to differentiate between scotomas that occur either due to perceptual issues or attentional issues. This would explain the incoherent picture that emerges from literature and textbooks.

### Interpretation

The results of the case study presented suggest that patients suffering from visual neglect could benefit from a virtual reality-based neuropsychological intervention based on restitutive concepts. Interestingly, the VFR intervention we used was not originally intended for neglect patients who usually suffer from lesions in higher cortical regions. The primary understanding of the scientific community and the clinical gold standard here is the use of so-called “compensatory procedures” (e.g., Kerkhoff et al. 2021). This means that in the course of training, patients are encouraged to make conscious eye movements into the blind field to improve awareness for this area. In contrast, “restitutive procedures” - as utilized in the SVFT in this study - rely on continuous fixation of a central point (usually in the form of a cross), thus forcing decentralized, peripheral awareness of the training stimuli displayed between defect and intact visual field. The background of this approach is - as mentioned above - the concept that these training stimuli reactivate the respective lesioned, topographically equivalent regions in early areas of the visual cortex. To date, we are not aware of any other study that has treated patients with neglect symptoms using an immersive procedure such as we did with virtual reality head mounted displays, combined with a restitutive procedure. Conversely, literature reviews show studies concentrating on visual scanning or pursuit trainings, neck-muscle vibration therapy, prism adaptation, visuo-motor feedback or other forms of attention training combined with magnetic or current stimulation (Kerkhoff, Rode & Clarke 2021; Pollock et al. 2019)

### Limitations

Our findings are limiting due to the nature of a case study, but concurrently indicate the potential of a virtual reality based neuropsychological intervention method that is based on a restitutive concept. Thus, at this point no generalized statements about the therapeutic effect size of the presented virtual reality training are valid. However, our findings warrant further and closer investigation (with a larger sample size) of the potential effective factors of the neuropsychological intervention that contributed to Patient 7 benefitting from an improved visual field both objectively and subjectively.

Another limitation of the present work relates to the functional improvement of patient 7’s daily life during/after therapy with the SVFT. In discussions after each assessment appointment, the patient’s general life situation and her coping with the visual field defect were discussed. However, Patient 7 did not report any conspicuous improvements in her daily life, although she consistently rated her visual impairment as improving over the training period. Thus, the present study does not provide structured and objectified data on the practical implications of Patient 7’s life situation during / after training with the SVFT. This area will also be further investigated and analyzed in continuing studies with larger numbers of participants.

## Conclusion

Our findings suggest that a virtual reality supported neuropsychological intervention based on restitutive concepts for patients suffering from visual neglect could improve visual field functionality and related visual perception after stroke or trauma.

## Data Availability

Data is available from the corresponding author upon reasonable request

## Acknowledgement

The authors sincerely thank Anja-Maria Ladek and Lydia Hell from the University Hospital Salzburg for Ophthalmology and Optometry for their medical and orthoptic support and Sarah Schuster for proof-reading and critical comment on the work.

## Ethics approval

The following study was reviewed and approved by the ethics commission of the University of Salzburg (Reference No. 39/2018)

## Contribution

All authors contributed equally to this work.

## Data availability statement

Data is available from the corresponding author upon reasonable request.

## References

Anderson A.J., Bedggood P.A., Kong Y.X.G., Martin K.R. & Yingrys A.J.V. (2017). Can Home Monitoring Allow Earlier Detection of Rapid Visual Field Progression in Glaucoma? Ophthalmology 124(12): 1735–1742. http://doi.org/10.1016/j.ophtha.2017.06.028

Azouvi P., Jacquin-Courtois S. & Luauté J. (2017). Rehabilitation of unilateral neglect: Evidence-based medicine. Annals of Physical and Rehabilitation Medicine 60(3): 191–197. https://doi.org/10.1016/j.rehab.2016.10.006

Balliet R., Blood K. & Bach-y-Rita P. (1985). Visual field rehabilitation in the cortically blind? Journal of Neurology Neurosurgery & Psychiatry 48: 1113–1124. https://doi.org/10.1136/jnnp.48.11.1113

Berti A., Garbarini F. & Neppi-Modona M. (2015). Chapter 32 - Disorders of Higher Cortical Function. Neurobiology of Brain Disorders. Biological Basis of Neurological and Psychiatric Disorders: 525–541. https://doi.org/10.1016/B978-0-12-398270-4.00032-X

Bisiach E. (1996). Unilateral Neglect and the Structure of Space Representation. Current Directions in Psychological Science 5(2): 62–65. https://doi.org/10.1111/1467-8721.ep10772737

Bouwmeester L., Heutink J. & Lucas C. (2007). The effect of visual training for patients with visual field defects due to brain damage: a systematic review. J Neurol Neurosurg Psychiatry 78: 555–564. https://doi.org/10.1136/jnnp.2006.103853

Bowen A., McKenna K. & Tallis R.C. (1999). Reasons for Variability in the Reported Rate of Occurrence of Unilateral Spatial Neglect After Stroke. Stroke 30(6): 1196–1202. https://doi.org/10.1161/01.STR.30.6.1196

Canning C.G., Allen N.E., Nackaerts E., Paul S.S., Nieuwboer A. & Gilat M. (2020). Virtual reality in research and rehabilitation of gait and balance in Parkinson disease. Nature Reviews Neurology 16: 409–425. https://doi.org/10.1038/s41582-020-0370-2

Chen P., Chen C.C., Hreha K., Goedert K.M. & Barrett A.M. (2015). Kessler Foundation Neglect Assessment Process Uniquely Measures Spatial Neglect During Activities of Daily Living. Archives of Physical Medicine and Rehabilitation 96(5): 869–876. https://doi.org/10.1016/j.apmr.2014.10.023

Cortés-Pérez I., Nieto-Escamez F.A. & Obrero-Gaitán E. (2020). Immersive Virtual Reality in Stroke Patients as a New Approach for Reducing Postural Disabilities and Falls Risk: A Case Series. Brain Sciences 10(5): 296–309. https://doi.org/10.3390/brainsci10050296

Demeyere N. & Gillebert C.R. (2019). Ego-and allocentric visuospatial neglect: Dissociations, prevalence, and laterality in acute stroke. Neuropsychology 33(4): 490–498. https://doi.org/10.1037/neu0000527

Frolov A., Feuerstein J. & Subramanian P.S. (2017). Homonymous Hemianopia and Vision Restoration Therapy. Neurologic Clinics 35: 29–43. https://doi.org/10.1016/j.ncl.2016.08.010

Glisson, C. (2006). Capturing the benefit of vision restoration therapy. Current Opinion in Ophthalmology 17: 504–508. https://doi.org/10.1097/icu.0b013e328010852e

Goh R.L.Z., Kong Y.X.G., McAlinden C., Liu J., Crowston J.G. & Skalicky S.E. (2018). Objective Assessment of Activity Limitation in Glaucoma with Smartphone Virtual Reality Goggles: A Pilot Study. Trans Vis Sci Tech 7(1): 10. https://doi.org/10.1167/tvst.7.1.10

Halligan P.W., Fink G.R., Marshall J.C. & Vallar G. (2003). Spatial cognition: evidence from visual neglect. Trends in Cognitive Sciences 7(3): 125–133. https://doi.org/10.1016/S1364-6613(03)00032-9

Horton J.C. (2005). Disappointing results from Nova Vision’s visual restoration therapy. British Journal of Ophthalmology 89: 1–2. https://doi.org/10.1136/bjo.2004.058214

Johnson C.A., Thapa S., Kong Y.X.G. & Robin A.L. (2017). Performance of an iPad Application To Detect Moderate and Advanced Visual Field Loss in Nepal. American Journal of Ophthalmology 182: 147–154. https://doi.org/10.1016/j.ajo.2017.08.007

Kerkhoff G., Rode G., Clarke S. Treating Neurovisual Deficits and Spatial Neglect. In: Platz T. (Ed.) Clinical Pathways in Stroke Rehabilitation. Evidence-based Clinical Practice Recommendations (2021). Cham. Springer. https://doi.org/10.1007/978-3-030-58505-1

Kerkhoff G. & Schenk T. (2012). Rehabilitation of neglect: An update. Neuropsychologia 50(6): 1072–1079. https://doi.org/10.1016/j.neuropsychologia.2012.01.024

Kolling G., Lachenmayr B., Vivell P. & Brandl H. (2012). Gutachterliche Prüfung der Sehfunktionen. In: Lachenmayr B. (eds) Begutachtung in der Augenheilkunde. Springer, Berlin, Heidelberg. https://doi.org/10.1007/978-3-642-29635-2_2

Leitner M.C., Guetlin D. & Hawelka S. (2021). Salzburg Visual Field Trainer (SVFT). A virtual reality device for (the evaluation of) neuropsychological rehabilitation. medRxiv. https://doi.org/10.1101/2021.03.25.21254352

Leitner M.C., Hutzler F., Schuster S., Vignali L., Marvan P., Reitsamer H. & Hawelka S. (2021). Eye-tracking-based visual field analysis (EFA): a reliable and precise perimetric methodology for the assessment of visual field defects. BMJ Open Ophthalmology, 6, e000429. https://doi.org/10.1136/bmjophth-2019-000429

Linden T., Samuelsson H., Skoog I. & Blomstrand C. (2005). Visual neglect and cognitive impairment in elderly patients late after stroke. Acta Neurologica Scandinavica 111(3): 163–168. https://doi.org/10.1111/j.1600-0404.2005.00391.x

Lindsay M.P., Norrving B., Sacco R.L., Brainin M., Hacke W., Martins S., Pandian J. & Feigin V. (2019). World Stroke Organization (WSO): Global Stroke Fact Sheet 2019. International Journal of Stroke 14(8): 806–817. https://doi.org/10.1177/1747493019881353

Lunven M. & Bartolomeo P. (2017). Attention and spatial cognition: Neural and anatomical substrates of visual neglect. Annals of Physical and Rehabilitation Medicine 60(3): 124–129. https://doi.org/10.1016/j.rehab.2016.01.004

Maggio M.G., De Luca R., Molonia F., Porcari B., Destro M., Casella C., Salvati R., Bramanti P. & Calabro R.S. (2019). Cognitive rehabilitation in patients with traumatic brain injury: A narrative review on the emerging use of virtual reality. Journal of Clinical Neuroscience 61: 1–4. https://doi.org/10.1016/j.jocn.2018.12.020

Marshall R.S., Chmayssani M., O’Brien K.A., Handy C., Greenstein V.C. (2010). Visual field expansion after visual restoration therapy. Clinical Rehabilitation 24: 1027–1035. https://doi.org/10.1177/0269215510362323

Matsumoto C., Yamao Y., Nomoto H., Takada S., Okuyama S., Kimura S., Yamanaka K., Aihara M. & Shimomura Y. (2016). Visual Field Testing with Head-Mounted Perimeter ‘imo’. PLoS ONE 11(8): e0161974. https://doi.org/10.1371/journal.pone.0161974

Mueller I., Mast H. & Sabel B.A. (2007). Recovery of visual field defects: A large clinical observational study using vision restoration therapy. Restorative Neurology and Neuroscience 25: 563-572. PMID: 18334773

Mueller I., Poggel D., Kenkel S., Kasten E. & Sabel B. (2003). Vision restoration therapy after brain damage: Subjective improvements of activities of daily life and their relationship to visual field enlargements. Visual Impairment Research, 5: 157–178. https://doi.org/10.1080/1388235039048692

Nesaratnam N., Thomas P.B.M., Kirollos R., Yingrys A.J., Kong G.Y.X. & Martin K.R. (2017). Tablets at the bedside - iPad based visual field test used in the diagnosis of Intrasellar Haemangiopericytoma: a case report. BMC Ophthalmology 17:53. https://doi.org/10.1186/s12886-017-0445-z

Poggel D., Kasten E. & Sabel B. (2004). Attentional cueing improves vision restoration therapy in patients with visual field loss. Neurology 6: 2069–2076. https://doi.org/10.1016/j.ajo.2005.02.012

Pollock A., Hazelton C., Rowe F.J, Jonuscheit S., Kernohan A., Angilley J., Henderson C.A., Langhorne P. & Campbell P. (2019). Interventions for visual field defects in people with stroke (Review). Cochrane Database of Systematic Reviews 5. https://doi.org/10.1002/14651858.CD008388.pub3

Qazi S., Raza K. (2020). Towards a VIREAL Platform: Virtual Reality in Cognitive and Behavioural Training for Autistic Individuals. In: Gupta D., Hassanien A., Khanna A. (eds) Advanced Computational Intelligence Techniques for Virtual Reality in Healthcare. Studies in Computational Intelligence, vol 875. Springer, Cham. https://doi.org/10.1007/978-3-030-35252-3_2

Reinhard J., Schreiber A., Schiefer U., Kasten E., Sabel B.A., Kenkel S., Vonthein R. & Trauzettel-Klosinski S. (2005). Does visual restitution training change absolute homonymous visual field defects? A fundus controlled study. British Journal of Ophthalmology 8: 30–35. https://doi.org/10.1136/bjo.2003.040543

Rosa A.M., Silva M.F., Ferreira S., Murta J. & Castelo-Branco M. (2013). Plasticity in the Human Visual Cortex: An Ophthalmology-Based Perspective. BioMed Research International, vol.2013. https://doi.org/10.1155/2013/568354

Rowe F.J., Hepworth L.R., Howard C., Hanna K.L., Cheyne C.P. & Currie J. (2019). High incidence and prevalence of visual problems after acute stroke: An epidemiology study with implications for service delivery. PLoS ONE 14(3): e0213035. https://doi.org/10.1371/journal.pone.0213035

Sabel B., Kasten E. & Kreutz M. (1997). Recovery of vision after partial visual system injury as a model of post-lesion neuroplasticity. Advances in Neurology 73: 251-276. PMID: 8959219

Sabel B., Kenkel S. & Kasten E. (2004). Vision restoration therapy (VRT) efficacy as assessed by comparative perimetric analysis and subjective questionnaires. Restorative Neurology and Neuroscience 2: 399-420. PMID: 15798360

Sabel B., Kenkel S. & Kasten E. (2005). Vision restoration therapy. British Journal of Ophthalmology 8: 522–524. https://doi.org/10.1136/bjo.2005.068163

Sabel B., Kruse R., Wolf F. & Guenther, T. (2013). Local topographic influences on vision restoration hot spots after brain damage. Restorative Neurology and Neuroscience 31: 787–803. https://doi.org/10.3233/RNN-139019

Sabel B. & Trauzettel-Klosinski, S. (2005). Improving vision in a patient with homonymous hemianopia. Journal of Neuroophthalmology 25: 143–149. https://doi.org/10.1097/01.wno.0000165107.24108.49

Sand K.M., Thomassen L., Naess H., Rodahl E. & Hoff J.M. (2012). Diagnosis and Rehabilitation of Visual Field Defects in Stroke Patients: A Retrospective Audit. Cerebrovasc Dis Extra 2:17–23. https://doi.org/10.1159/000337016

Schreiber A., Vonthein R., Reinhard J., Trauzettel-Klosinski S., Connert C. & Schiefer U. (2006). Effect of visual restitution training on absolute homonymous scotomas. Neurology 67: 143–145. https://doi.org/10.1212/01.wnl.0000223338.26040.fb

Schulz A.M., Graham E.C., You Y., Klistorner A. & Graham S.L. (2017). Performance of iPad-based threshold perimetry in glaucoma and controls. Clinical & Experimental Ophthalmology 46: 346–355. https://doi.org/10.1111/ceo.13082

Spofforth J., Codina D. & Bjerre A. (2017). Is the ‘Visual Fields Easy’ Application a Useful Tool to Identify Visual Field Defects in Patients Who Have Suffered a Stroke?. Ophthalmology Research: An International Journal 7(1): 1–10. https://doi.org/10.9734/OR/2017/34947

Spreij L.A., Visser-Meily J.M.A., Sibbel J., Gosselt I.K. & Nijboer T.C.W. (2020). Feasibility and user-experience of virtual reality in neuropsychological assessment following stroke. Neuropsychological Rehabilitation. https://doi.org/10.1080/09602011.2020.1831935

Ten Brink A.F., Verwer J.H., Biesbroek J.M., Visser-Meily J.M.A. & Nijboer T.C.W. (2016). Differences between left-and right-sided neglect revisited: A large cohort study across multiple domains. Journal of Clinical and Experimental Neuropsychology 39 (7): 707–723. https://doi.org/10.1080/13803395.2016.1262333

Trauzettel-Klosinski (2011). Current methods of visual rehabilitation. Dtsch Arztebl Int 108(51-52): 871–878. https://doi.org/10.3238/arztebl.2011.0871

Umarova R.M., Saur D., Kaller C.P., Vry M.S., Glauche V., Mader I., Hennig J. & Weiller C. (2011). Acute visual neglect and extinction: distinct functional state of the visuospatial attention system. Brain 134(11): 3310–3325. https://doi.org/10.1093/brain/awr220

Wroblewski D., Francis B.A., Sadun A., Vakili G. & Chopra V. (2014). Testing of Visual Field with Virtual Reality Goggles in Manual and Visual Grasp Modes. BioMed Research International. https://doi.org/10.1155/2014/206082

Zihl J. & von Cranon D. (1985). Visual field recovery from scotoma in patients with postgeniculate damage. A review of 55 cases. Brain 108: 335–365. https://doi.org/10.1093/brain/108.2.335

